# Centromedian thalamic deep brain stimulation for idiopathic generalized epilepsy – case series and target analysis

**DOI:** 10.1101/2024.02.26.24303317

**Authors:** Sihyeong Park, Shruti Agashe, Gamaleldin Osman, Hugh D. Simpson, Kai J. Miller, Jamie J. Van Gompel, Keith Starnes, Brian N. Lundstrom, Gregory A. Worrell, Nicholas M. Gregg

## Abstract

**Introduction:** There is a lack of treatment options for individuals with drug resistant idiopathic generalized epilepsy (IGE). Small, limited case series suggest that centromedian thalamus deep brain stimulation (CM-DBS) may be an effective treatment option. The optimal CM-DBS target for IGE is underexamined. Here, we present a retrospective analysis of CM-DBS targeting and efficacy for five patients with drug-resistant IGE.

**Methods:** This single-center case series study included all patients with drug resistant IGE treated with CM-DBS to date. Seizure outcomes were measured with patient-reported seizure frequency. Baseline frequency was taken from the visit immediately before implantation, and it was compared with the seizure frequency from the most recent visit. Stimulation parameters from last clinic visit were used to determine the volume of tissue activated (VTA) by DBS. Lead proximity to target, and VTA overlap with CM nucleus was performed using an open-source toolbox (Lead-DBS 3.0, SimBio/FieldTrip). Associations between the intersection of the VTA and CM nucleus, and seizure outcomes were measured using Spearman’s rank correlation.

**Results:** Five patients were identified and included in the study. Median age was 31 (22-45). Median baseline frequency of convulsive seizure was 1.5 per month (0-3.5). Median follow-up time was 13 months (3-23). Median convulsive seizure frequency reduction was 60% (−200-100%). One patient had only absence seizures, with >99% absence seizure frequency reduction. Four out of five subjects had electrode contacts positioned within the CM nucleus target, all of whom had >50% reduction in primary semiology seizure frequency (3 subjects with convulsive seizures, 1 patient with absence seizures). Spearman’s rho was 0.90 (P=0.025). Volumetric “sweet-spot” mapping revealed best outcomes were correlated with stimulation of the middle ventral CM nucleus.

**Conclusion:** Our findings suggest that CM-DBS can be an effective treatment for patients with IGE. The extent of overlap between the VTA by DBS and CM nucleus is correlated with the degree of seizure reduction, highlighting the importance of accurate targeting and volumetric targeting analysis.

## 1. Introduction

Idiopathic generalized epilepsy (IGE) consists of childhood absence epilepsy, juvenile absence epilepsy, juvenile myoclonic epilepsy, and epilepsy with generalized tonic clonic (GTC) seizures alone^1^. Incidence of IGE is reported to be 2.7/100 000 person-years^2^, and a recent meta-analytic study described that 24% of patients with IGE were drug-resistant^3^. Treatment options for drug resistant IGE are lacking, and there are no approved neuromodulation therapies with an indication for IGE in the US.

Small, limited case series suggest that centromedian thalamus deep brain stimulation (CM-DBS) may be an effective treatment option for drug resistant IGE. Study groups included in previous investigations were often heterogeneous, grouping individuals with a spectrum of genetic generalized epilepsies and developmental and epileptic encephalopathies^4^. Analysis of data from a prospective randomized controlled trial of CM-DBS for Lennox-Gastaut Syndrome (LGS) suggests that efficacy is associated with stimulation of the parvocellular CM and ventral lateral nucleus of the thalamus, while optimal targeting of CM-DBS for IGE remains underexamined. Furthermore, prior work comparing interictal epileptiform discharges in individuals with LGS and IGE demonstrates distinct patterns of cortical activation/inhibition between these groups, suggesting that optimal neuromodulation targets and paradigms may be specific to the epilepsy syndrome—perhaps a validation for splitters over lumpers^5^. Here, we present a retrospective analysis of CM-DBS targeting and efficacy for five patients with drug-resistant IGE.

## 2. Methods

### 2.1. Study design

This single-center case series study included all patients with drug resistant IGE treated with CM-DBS to date at our institution. Patients were included consecutively, and retrospective review of the electronic medical record was performed to obtain outcome variables. Data from subject 2 has been previously published^6^. This study was approved by the Mayo Clinic Institutional Review Board.

### 2.2. Variables

Seizure outcomes were measured with patient-reported seizure diaries. Baseline frequency was taken from the visit immediately before the implantation, and it was compared with the seizure frequency from the most recent clinic visit. A positive percent reduction of seizure frequency indicates a lower seizure frequency at the last visit compared to the baseline seizure frequency. A negative percent reduction of seizure frequency implies a higher, worsening, seizure frequency. Unprovoked seizures were included for analyses. Patient 2 had a cluster of seizures following abrupt discontinuation of clonazepam, and these seizures were not included—subsequently, patient 2 remained seizure-free for 7 months following reinstitution of clonazepam, through last follow up.

### 2.3. Targeting, lead localization, and VTA modeling

Indirect targeting, performed based on anterior commissure (AC)-posterior commissure (PC) offset, was applied to patient #1. Subsequent implants were conducted after the implementation of thalamus atlas warp-based targeting using the Krauth/Morel atlas^7^, similar to prior work^6^.

DBS lead localization and modeling of the volume of tissue activated (VTA) by DBS were performed as previously described^6^ using open-source Lead-DBS toolbox version 3.0^8^. Briefly, postoperative CT was co-registered to preoperative MRI, images were spatially normalized into Montreal Neurological Institute (MNI) space, and DBS electrodes were localized in patient and MNI space using the PaCER algorithm. DBS electrode localizations were corrected for brain shift by affine transform restricted to a subcortical area of interest. Thalamic nuclei were represented using the Krauth/Morel atlas^7^.

### 2.4. Probabilistic mapping

Sweet-spot analysis was performed using Lead-DBS toolbox. To maximize the statistical power, all electrodes were warped to the right hemisphere. The VTAs were weighted by patient’s percent reduction of seizure frequency. Voxel threshold of 60% was applied, such that only the voxels covered by the VTA from more than 60% of the leads were included in the analysis. Subsequently, t-test was performed at each voxel, with the null hypothesis equal to the mean seizure reduction across the cohort.

### 2.5. Statistical analysis

Responder was defined as a patient with a reduction of seizure frequency by 50% or greater. Associations between the intersection of the VTA and CM, and seizure outcomes were measured using Spearman’s rank correlation.

## 3. Results

### 3.1. Patient demographics

Five patients were identified and included in the study (Table 1). Median age was 31 (range: 22-45). Eighty percent of the patients were female. Median number of the antiseizure medications (ASMs) the patient had tried was 7 (range: 4-10), and the median number of the current ASMs was 3 (range: 2-4). During follow-up, one patient (patient #3) had an addition of an ASM, while two patients (patient #2, 4) underwent ASM reduction. The remaining patients had no changes in ASM regimen.

**Table 1.** Sumary of patient characteristics and results. Bolded are the primary seizure type. CBD, cannabidiol; CLB, clobazam; CZP, clonazepam; FBM, felbamate; GAB, gabapentin; IPG, implantable pulse generator; KD, ketogenic diet; LEV, levetiracetam; LCM, lacosamide; LTG, lamotrigine; mo, months; mKD, modified ketogenic diet; sz, seizure; OXC, oxcarbazepine; PHT, phenytoin; PRP, perampanel; RUF, rufinamide; TPM, topiramate; VGB, vigabatrin; VNS, vagus nerve stimulator; VPA, valproic acid; ZNS, zonisamide

| Patient/IPG | Electrode model | Age | Gender | Epilepsy onset | Semiology | Follow-up time | ASDs trialed pre-DBS; ASD at time of implant | Stimulation settings at last visit (frequency, pulse width, amp) | Stimulation parameters |  | Convulsive seizure frequency (sz/mo) |  | GTCSreduction (%) | Absence reduction (%) | Myoclonic reduction (%) |
| --- | --- | --- | --- | --- | --- | --- | --- | --- | --- | --- | --- | --- | --- | --- | --- |
|  |  |  |  |  |  |  |  |  | Right CM | Left CM | GTCSpre | GTCSpost |  |  |  |
| 1/Medtronic. Activa PC | Medtronic model 3387 | 40-44 | F | 0-9 yo | Abs, Myo, **GTCS** | 7 mo | VPA, PRP, LCM, LTG, BRV, LEV, CBZ, PHB, TPM, PHT → FEL, ZNS, VNS | 60 Hz, 90 usec, 2 V | 8o | 0o | 1 | 3 | -200 | . | . |
|  |  |  |  |  |  |  |  |  | 9- | 1- |  |  |  |  |  |
|  |  |  |  |  |  |  |  |  | 10- | 2- |  |  |  |  |  |
|  |  |  |  |  |  |  |  |  | 11o | 3o |  |  |  |  |  |
| 2/Medtronic. Activa PC | Medtronic model 3387 | 30-34 | F | 0-9 yo | Abs, **GTCS** | 13 mo | OXC, VAL, ETX, LCM, LEV, TPM → LTG, ZNS, BRV, CZP, mKD, VNS | 60 Hz, 90 usec, 4.5V | 8- | 0- | 1.5 | 0 | 100 | . | N/A |
|  |  |  |  |  |  |  |  |  | 9o | 1o |  |  |  |  |  |
|  |  |  |  |  |  |  |  |  | 10o | 2o |  |  |  |  |  |
|  |  |  |  |  |  |  |  |  | 11o | 3o |  |  |  |  |  |
| 3/Medtronic. Percept PC. B35200 | Medtronic SenSight directional leads B33015-42 | 20-24 | F | 10-19 yo | Abs, Myo, **GTCS** | 23 mo | PRP, VPA, LEV, TPM, ZNS, CLB, CBD, KD → BVA, CZP, ETX, LTG, VNS | 7Hz, 200 usec, 4.0 mA | 8o | 0o | 2.5 | 1 | 60 | 75 | 75 |
|  |  |  |  |  |  |  |  |  | 9o | 1o |  |  |  |  |  |
|  |  |  |  |  |  |  |  |  | 10- | 2- |  |  |  |  |  |
|  |  |  |  |  |  |  |  |  | 11o | 3o |  |  |  |  |  |
| 4/Medtronic. Percept PC | Medtronic SenSight B3300542M | 30-34 | Non-binary | 10-19 yo | Abs, Myo, **GTCS** | 21 mo | VPA, LTG, PRP, RUF, FBM, TPM, LEV, ETX, ZNS, CBD → LCM, BRV, CLB | 60 Hz, 90 usec, 4.0 mA | 8o | 0- | 3.5 | 1 | 71.43 | None at baseline | . |
|  |  |  |  |  |  |  |  |  | 9- | 1o |  |  |  |  |  |
|  |  |  |  |  |  |  |  |  | 10o | 2o |  |  |  |  |  |
|  |  |  |  |  |  |  |  |  | 11o | 3o |  |  |  |  |  |
| 5/Boston Scientific. M365DB12160. Vercise™ Genus R16 IPG. | Vercise Cartesia Directional leads DB-2202-45 | 45-49 | F | 10-19 yo | **Abs**, **GTCS** | 3 mo | LEV, PHT, VPA, GAB, LTG, CBD, ETX → TPM, LCM | 60 Hz, 90 usec, 3.0 mA | Lv 1o | Lv 1- | 0 | 0 | 0 | >99% | N/A |
|  |  |  |  |  |  |  |  |  | Lv 2- | Lv 2- |  |  |  |  |  |
|  |  |  |  |  |  |  |  |  | Lv 3- | Lv 3o |  |  |  |  |  |
|  |  |  |  |  |  |  |  |  | Lv 4o | Lv 4o |  |  |  |  |  |

At the time of the last clinic visit, 4 patients were receiving 60 Hz stimulation (90 μsec pulse width). One patient was receiving low-frequency (7 Hz) stimulation after failing 55 Hz stimulation (90 μsec pulse width, left lead: 2a: 0.2mA, 2b: 0.2mA, 2c: 1.3mA, right lead 9a: 0.4mA, 9b: 0.4mA, 9c: 0.4mA). All were on continuous stimulation settings.

### 3.2. CM-DBS outcome

Median baseline frequency of convulsive seizures was 1.5 per month (range: 0-3.5 per month). Median follow-up time was 13 months (range: 3-23 months). Median convulsive seizure frequency reduction was 60% (range: -200-100%). Four patients had active contacts positioned within the CM nucleus target, all of whom had had >50% reduction in primary semiology seizure frequency (3 patients with GTC seizures, 1 patient with absence seizures). The median percent reduction of GTC seizure for these four patients was 66% (Figure 1). One of these four patients (patient #5) had no GTC seizures at baseline—her dominant seizure type was absence seizure, with 30-40 absence seizures daily at baseline, confirmed by epilepsy monitoring unit overnight video EEG monitoring on home medications. She had a remarkable response, reporting 5 seizures per month with CM-DBS, a greater than 99% reduction in seizure frequency, on a stable ASM regimen. Patient 1 did not have active contacts positioned within the CM nucleus and did not respond to CM-DBS, and experienced an increase in seizure frequency with stimulation and head and neck pain. This patient had device explantation 7 months post implant.

**Figure 1.**
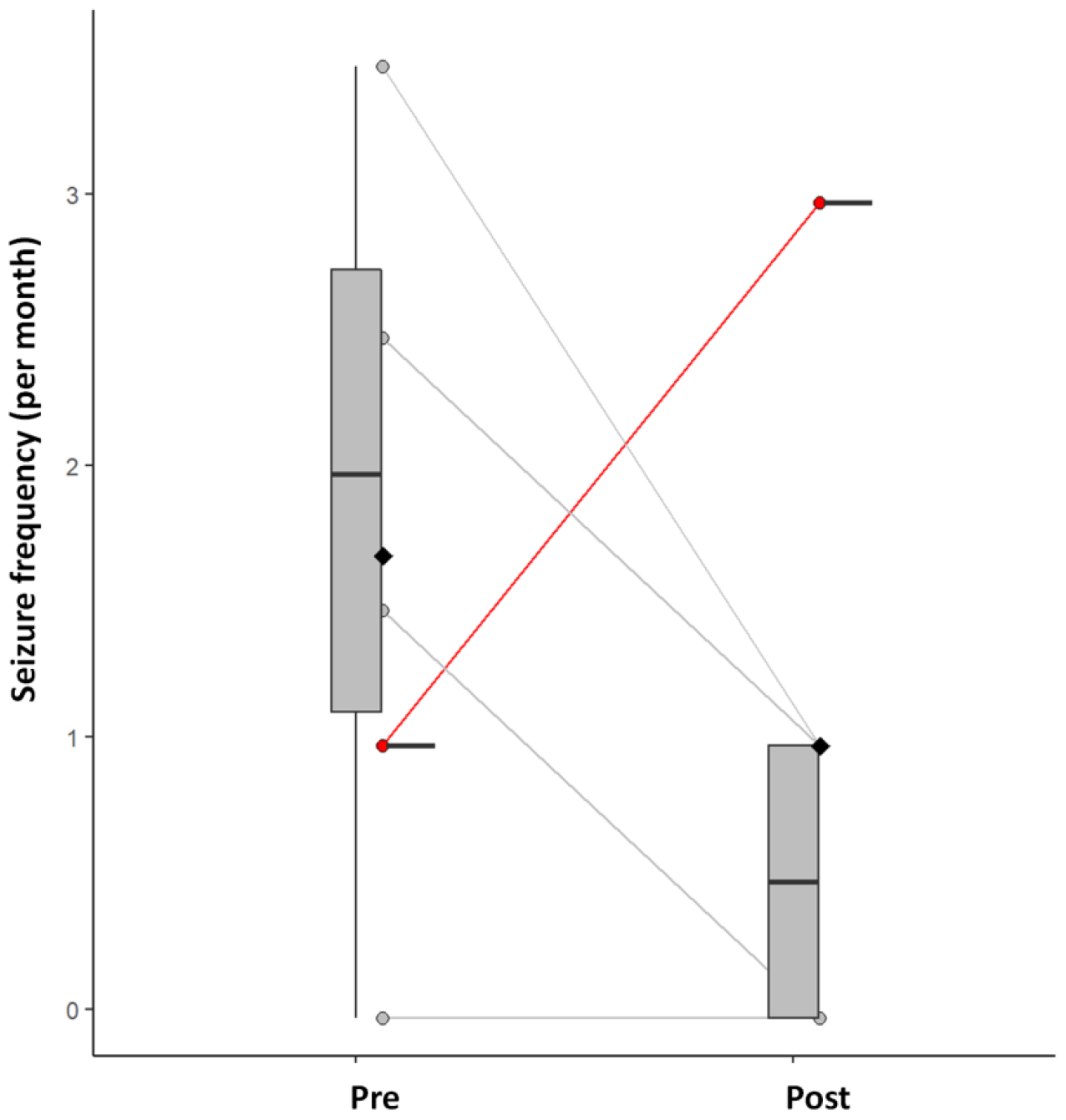
Boxplot shows generalized tonic-clonic seizure frequency (left)) pre- and post-DBS in responders (grey) and non-responder (red) (all responders had active electrodes positioned within the CM nucleus; the non-responder did not have electrodes positioned within the CM nucleus).

### 3.3. Target analysis and volumetric mapping

Figure 2 shows the group rendering of the electrodes relative to the centromedian-parafascicular (CM-Pf) complex. The electrodes of the responders and their active contacts (red electrode contacts on the grey-colored responder leads) were positioned within the CM-Pf target, whereas the single non-responder (yellow-colored leads) had electrode placement posteromedial to the CM nucleus. This patient was implanted prior to implementation of individualized atlas-based targeting.

**Figure 2.**
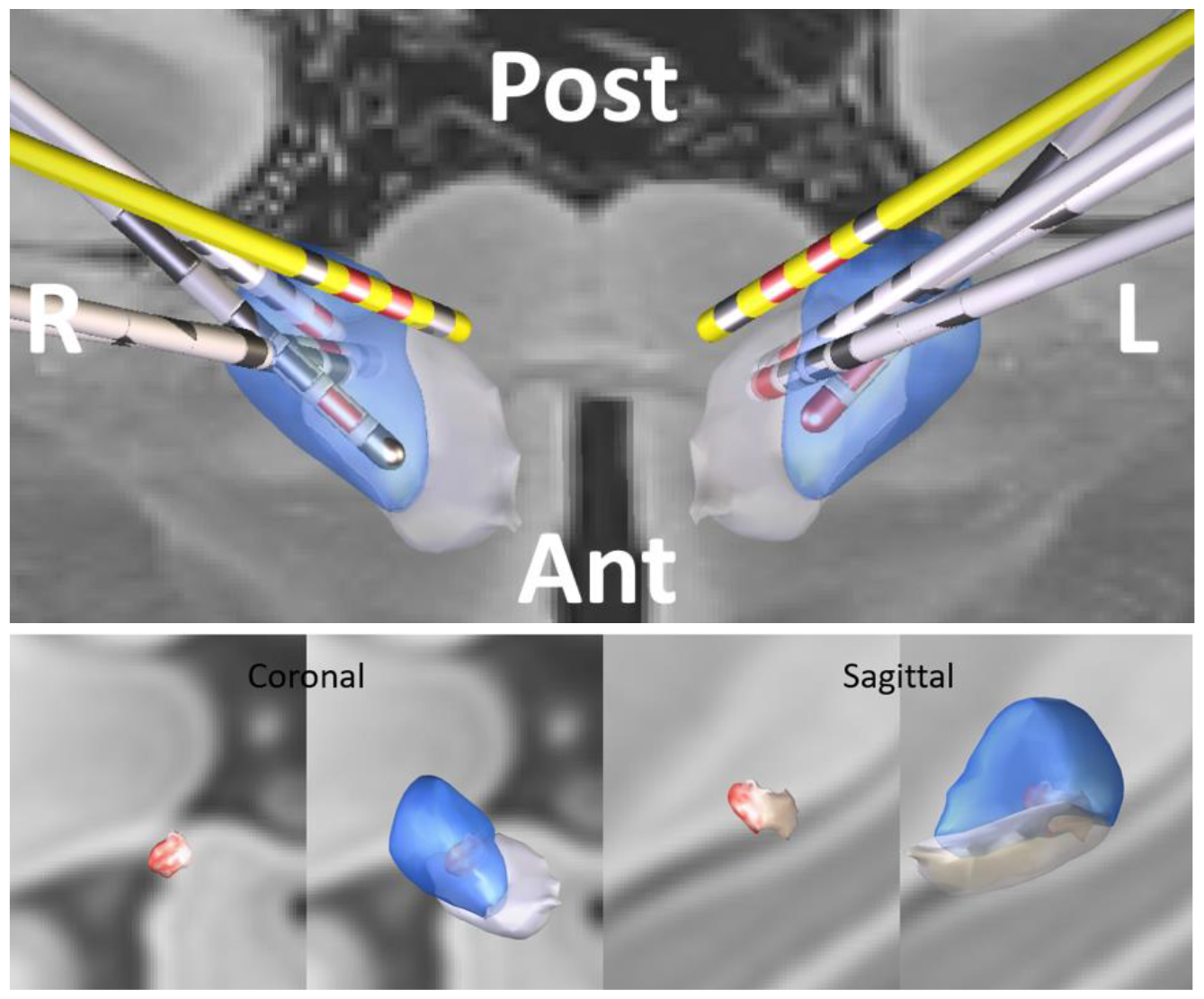
Top panel: CM-DBS leads for 5 subjects with IGE; responder leads (grey), non-responder leads (yellow). Bottom Panels: All VTAs were warped into the right hemisphere for “sweet-spot” analysis. Red voxels indicate regions with greater seizure reduction outcome. Centromedian nucleus (blue); parafascicular nucleus (grey).

Three-dimensional rendering of the VTA for each responder showed substantial overlap with the CM nucleus (Figure 3), with median VTA-to-CM overlap (i.e. CM:VTA intersection) of 72.76 mm3 (range: 38.25-208.95 mm^3^). Spearman’s rho rank correlation comparing CM:VTA intersection volume to the seizure frequency reduction was 0.90 (P=0.025). Volumetric sweet-spot mapping revealed best outcomes with stimulation of the middle ventral CM nucleus.

**Figure 3.**
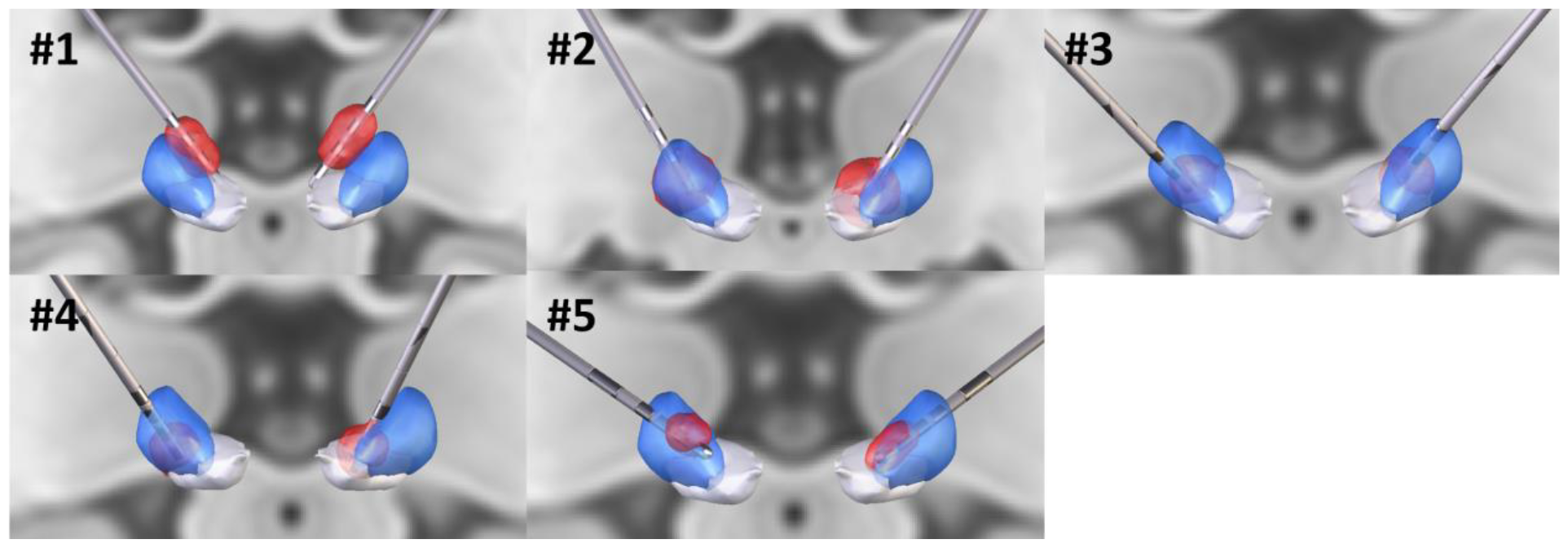
Volume of tissue activated (VTA) of individual patients. #1: non-responder. #2-5: responders.

## 4. Discussion

Our study shows excellent efficacy of CM-DBS in patients with drug-resistant IGE with well positioned electrodes in the CM nucleus, with 80% overall responder rate, and 100% responder rate with well positioned leads (4 out of 5 subjects). Larger CM-VTA intersection, reflecting accurate targeting, was significantly correlated with greater seizure control. Patient 1 did not respond to CM-DBS, which may relate to suboptimal electrode position (no leads positioned within the CM target). Subject 1 did not have CM-atlas based targeting, in contrast to the remainder of the cohort, underscoring the importance of new individualized approaches to atlas-based targeting^9^ and direct targeting with novel MRI sequences (MP2RAGE)^10^. In addition, this warrants implementation of volumetric targeting into a routine practice and underlines importance of describing targeting in DBS outcome studies.

Our results suggest that the stimulation of the middle ventral aspect of the CM was associated with better clinical outcome in our 5 person cohort. This finding is in contrast to a recent randomized controlled trial of CM-DBS for Lennox-Gastaut syndrome (ESTEL trial), with optimal stimulation targeting the parvocellular CM and ventral lateral nucleus of the thalamus^11^. The ESTEL trial had a reported responder rate of 50% and median seizure frequency reduction of 47%.^12^ A tractography-based study^13^ of the ESTEL trial cohort demonstrated stronger connectivity between the sweet-spot and cerebellum as well as frontal lobes. Prior working using simultaneous EEG with functional MRI has demonstrated distinct activation patterns between LGS and IGE, with increased BOLD signal in frontoparietal area in LGS and in sensorimotor cortex in IGE^5^. Distinct seizure networks may result in syndrome-specific optimal stimulation targets.

In addition to CM-DBS, efficacy of CM-responsive neurostimulation (RNS) in drug-resistant IGE has been studied. A four subject case series reported 75-99% reduction in seizure frequency with CM-RNS^14^. A multicenter randomized study on CM-RNS is ongoing (ClinicalTrials.gov ID: NCT05147571). The results of this study may help optimize CM-DBS, including targeting and stimulation parameters. Establishing the efficacy of non-responsive options such as DBS will provide options for patients in addition to data mounting for closed loop options such as RNS making this an important experience.

This study is limited by the relatively small number of individuals with drug resistant IGE treated with CM-DBS. Further studies with large sample size and more varied electrode targeting are needed to evaluate the relative performance of differing “sweet spots” reported in the literature. Our study is limited to individuals with IGE, which may in part explain differences in results relative to prior studies that included individuals with diverse genetic generalized epilepsy and developmental and epileptic encephalopathy syndromes. Prior literature supports efficacy of low frequency stimulation in thalamic deep brain stimulation^15^. Further work is needed to investigate the effectiveness of different stimulation parameters (high vs. low frequency stimulation)—one subject here responded to low frequency but not her frequency stimulation—and to better study outcomes over time.

## 5. Conclusion

Our findings support that non-responsive, continuous CM-DBS as an effective treatment for patients with drug-resistant IGE. The volume of intersection between the VTA and CM nucleus is correlated with the degree of seizure reduction, highlighting the importance of accurate targeting. Further work, with larger cohorts, and more varied electrode targeting are required to establish the relative performance between different “sweet spots” described in the literature.

## Data Availability

Data will be made available upon reasonable request.

## Author Contributions

N.G. conceptualized the study. S.P. and N.G. designed the study. S.P. obtained the data and performed target analysis, volumetric mapping, and statistical analysis. S.P. and N.G. drafted the manuscript. S.P., S.A., G.O., H.D.S., K.M., J.V.G. K.S., B.L., G.W., and N.G. provided critical feedback to the manuscript.

## Acknowledgement

None

## Conflict of Interest

G.A.W., B.N.L., J.J.V.V., declare intellectual property licensed to Cadence Neuroscience (BNL waived contractual rights). S.A. is a consultant for Blackrock Neurotech. B.N.L. declares intellectual property licensed to Seer Medical (contractual rights waived). G.A.W. licensed intellectual property and serves on the scientific advisor board of NeuroOne, Inc. B.N.L., G.A.W, N.M.G. are investigators for the Medtronic Deep Brain Stimulation Therapy for Epilepsy PostApproval Study. B.N.L is an investigator for the Neuroelectrics tDCS for Patients with Epilepsy Study. J.J.V.V., G.A.W., B.N.L., N.M.G. are investigators for the NeuroPace RNS NAUTILUS study. N.M.G has consulted for NeuroOne, Inc. (funds to Mayo Clinic). B.N.L has consulted for Epiminder, Medtronic, Neuropace, and Philips Neuro (all funds to Mayo Clinic). The remaining authors declare no competing interests.

This work was approved by the Mayo Clinic Institutional Review Board. Data will be made available upon reasonable request.

## Notes

### Funding Statement

This study did not receive any funding.

### Author Declarations

IRB of Mayo Clinic gave ethical approval for this work (ID: 23-009255)

